# Causal evidence linking chronic pain genetics to late-onset asthma via the nervous system

**DOI:** 10.1101/2025.08.08.25333309

**Authors:** Goodarz Kolifarhood, Marc Parisien, Matt Fillingim, Charlise Chen, Yiran Chen, Nikolay Dimitrov, Maha Zidan, Lovéni Hanumunthadu, Peter Hyunwuk Her, Sahir Bhatnagar, Luda Diatchenko, Audrey V. Grant

**Affiliations:** Department of Anesthesia, Faculty of Medicine and Health Sciences, McGill University Montreal, Quebec, Canada; Faculty of Dental Medicine and Oral Health, McGill University, Montreal, Quebec, Canada; Alan Edwards Centre for Research on Pain, McGill University, Montreal, Quebec, Canada; Institut de recherches cliniques de Montréal (IRCM), Montreal, Quebec, Canada; Faculty of Engineering, McGill University, Montreal, Quebec, Canada; Department of Medical Biophysics, University of Toronto, Toronto, Ontario, Canada; Princess Margaret Cancer Center, University Health Network, Toronto, Ontario, Canada; Department of Epidemiology, Biostatistics and Occupational Health, Faculty of Medicine and Health Sciences, McGill University Montreal, Quebec, Canada

**Keywords:** Chronic Pain, Asthma, Aging, Genetics, Genomics, Expression, Comorbidity, Longitudinal, Polygenic Risk Score, Genomewide Association

## Abstract

**Background:** Chronic pain and asthma are associated, but the direction and basis of their genetic and biological relationship remain unclear.

**Methods:** We conducted genome-wide association (GWA), cross-trait meta-analysis, polygenic risk score (PRS) prediction, bivariate causal modeling, and Mendelian randomization (MR) across nine chronic pain traits and three asthma age-of-onset strata (<18, 18–40, and >40 years for childhood-, adult-, and late-onset asthma) in 456,958 UK Biobank (UKB) and 25,275 Canadian Longitudinal Study on Aging (CLSA) participants of European descent. We analyzed shared and distinct genetic architecture using gene-, pathway-, tissue-, and cell-type-based enrichment analyses.

**Results:** Multisite chronic pain (MCP) showed the strongest and most consistent genetic overlap with asthma, with genetic correlation increasing from childhood (rg = 0.01) to late-onset asthma (rg = 0.40). Estimated causal variants for late-onset asthma (∼1.8 K) were nested within a broader MCP profile (∼9.4 K), with fewer for childhood asthma (∼0.2 K). Using PRS, MR, and longitudinal analyses, we found that MCP contributes causally to late-onset asthma. Top causal variants from MR mapped to *GMPPB–RNF123*, *DCC*, and *FOXP2*. Conditioning by MCP amplified late-onset asthma variant effect sizes and uncovered genes enriched for immune and central nervous system pathways, tissues, and cell types. In contrast, childhood asthma showed immune-specific enrichment alone.

**Conclusion:** These findings reveal neurological function linking chronic pain to late-onset asthma, distinct from childhood asthma, and highlight a central nervous system contribution to asthma emerging later in life.

## Introduction

Across the lifespan, chronic pain conditions often emerge and intensify in middle to older age, while asthma onset can occur at any age. Chronic pain, defined as pain lasting three or more months without substantial tissue damage, affects approximately 20% of the global population and half of older adults leading to significant individual and societal impacts^1,2^. Chronic pain is frequently accompanied by other clinical conditions such as depression, sleep disorders, cardiovascular diseases, asthma, and inflammatory diseases, further exacerbating quality of life^3,4^. Similarly, asthma is a complex, chronic disorder. It is characterized by bronchial inflammation and narrowing of the airways, with prevalence rates ranging from 1% to 18% worldwide depending on demographic and environmental factors^5,6^. Asthma varies in inflammatory profiles, severity, allergic or non-allergic forms, and has distinct etiological features for childhood versus adult-onset^7–9^.

These two seemingly disparate conditions share inflammation as a key pathophysiological mechanism. Furthermore, reciprocal interactions between the nervous and immune systems play a central role in the initiation and persistence of both chronic pain and asthma^10,11^. In chronic pain, pro-inflammatory cytokines involved in regulating neurons and glial cells can trigger the transmission of nociceptive signals to the brain^12^. In asthma, exposure to external stimuli activates dendritic cells, leading to the secretion of pro-inflammatory cytokines that interact with sensory nerves, causing the release of neuropeptides and amplifying inflammation^13^. Moreover, emerging evidence suggests an interface connecting cell-types implicated in pain and asthma, notably the activation of sensory neurons initiates airway inflammation by releasing mediators from parasympathetic neurons through smooth muscle contraction^14^. Additionally, silencing nociceptor neurons reduces allergic airway inflammation in animal models^15^.

Genome-wide association (GWA) studies for chronic pain have highlighted neurological pathways and functions involved in neurotransmission, pain perception and development of the nervous system^16–18^, while for asthma, they have identified immune pathways and functions associated with inflammatory responses and airway regulation^19–21^. Some GWA studies of chronic pain conditions have also pointed to immune system modulation and inflammation pathways including B-cell involvement^17,22–24^. A cross-trait chronic pain approach pointed to immune system function associated with chemokine secretion and T-cell function^25^. In considering the genetics of chronic pain and asthma, a common immune rather than neurological component underlying the conditions seems the most likely based on current knowledge.

Chronic pain and asthma are genetically correlated, with a UK Biobank study reporting 20% overlap between multisite chronic pain (MCP) and asthma¹ . However, it remains unclear how these conditions are biologically linked, and whether chronic pain contributes causally to asthma—or vice versa. We therefore used an integrative framework to disentangle shared and distinct genetic architecture across chronic pain traits and asthma stratified by age of onset.

## Materials and methods

### Discovery study cohort: UKB

This study leveraged data from the UK Biobank (UKB), a large-scale population cohort of individuals aged 37–73 years across the United Kingdom. Participants were recruited voluntarily between 2006 and 2010, with baseline and first follow-up data collected through questionnaires (socio-demographic, lifestyle, and health), physical measurements, and blood samples^26^.

### Chronic pain, asthma, and COPD status

Chronic pain phenotypes were determined based on pain reports at specific body sites—back, neck and shoulder, hip, knee, stomach and abdomen, headache, facial, and pain over body (widespread). A musculoskeletal pain phenotype was defined based on back, neck and shoulder, hip, and knee pain. Multisite chronic pain (MCP), a quantitative trait, reflects the count of body sites with a report of chronic pain. Doctor-diagnosed asthma was identified via self-reported non-cancer illnesses and stratified by age of onset; <18 years (childhood, *N*=15,063),18-40 years (adult, *N*=13,970), and > 40 years (late, *N*=19,764) to capture heterogeneity in disease presentation. Chronic Obstructive Pulmonary Disease (COPD) was included to account for potential misclassification of asthma cases and its influence on epidemiological and genetic associations with chronic pain (**see Supplementary Materials**).

### Identification of European ancestry individuals (EUR)

To identify EUR, we applied a standard genetic clustering method, expanding beyond UKB’s “White British” label^27^. Participants with sex discrepancies or who opted out of the study were excluded, resulting in 456,958 individuals (**see Supplementary Materials**).

### Phenotypic associations

Phenotypic associations between age-of-onset-stratified asthma and chronic pain traits were assessed using generalized linear models in R, adjusting for sex and age, within a random subset of UKB participants (80% of cases, 90% of controls) used for PRS training; the remainder were used for PRS testing. (**see Supplementary Materials**).

### Genome-wide association scans

Genome-wide association (GWA) scans were conducted using Regenie (v3.1.3)^28^. Covariates included age, sex, and the top 40 principal components. Post-GWA scan filtering excluded variants with low frequency (MAF < 0.01), low imputation quality (Info score ≤ 0.3), and Hardy–Weinberg disequilibrium (*P* < 10^−6^). A total of 9.4 million variants were retained for analysis. Lead SNPs were identified through the online platform for functional mapping and annotation of GWA studies (FUMA), using linkage disequilibrium (LD) thresholds to define independent loci. Functional annotations and gene mappings were performed using ANNOVAR and Ensemble data^29^(**see Supplementary Materials**).

### Heritability, genetic correlation, and shared genetic architecture

Heritability and genetic correlations were estimated using LDSC^30^. Shared or trait-specific polygenic architecture was examined with MiXeR. MiXeR modeled discoverability (proportion of variance detectable at genome-wide significance) and polygenicity (number of variants explaining 90% of heritability) under a bivariate model across asthma and pain traits to estimate number of trait-specific or overlapping causal genetic variants^31,32^(**see Supplementary Materials**).

### Meta-analyses

To increase statistical power, we conducted trait-specific meta-analyses using MTAG using asthma or chronic pain traits as the primary trait, and traits from the other category as the secondary trait, which accounts for overlapping samples and adjusts for LD structure. MTAG-enhanced summary statistics were then used to refine gene prioritization^33^(**see Supplementary Materials**).

### Polygenic risk score

Polygenic scores were constructed for MCP and asthma traits using PRSice^34^. Scores were validated in independent UKB samples (20% of cases and 10% of controls who were not included in primary GWAS) and externally in 25,275 samples with European ancestry in the Canadian Longitudinal Study on Aging (CLSA)^35,36^, testing cross-phenotype prediction (**see Supplementary Materials**).

### Mendelian randomization and longitudinal analyses

Mendelian Randomization (MR) using CAUSE was conducted to assess causal and pleiotropic relationships between traits, accounting for LD and sample overlap by incorporating all variants^37^. To support the MR findings, longitudinal analyses using logistic regression were performed to examine directional associations between asthma age-of-onset and chronic pain across baseline and follow-up data (**see Supplementary Materials**).

### Biological function analyses

We conducted gene-, gene set–, and gene-property–level analyses using MAGMA (v1.08) on both primary and MTAG-boosted GWAS results^38^. Gene-level associations were tested using SNP-wise models to compute gene-based p-values. Tissue and cell-type enrichment analyses were performed using expression data from GTEx and the Human Protein Atlas^39–42^, and cell-type specificity was assessed via ct-LDSC across multiple transcriptomic datasets^43,44^. Additionally, CAUSE-derived ELPD values were used to identify causal and pleiotropic pathways through SNP-to-gene mapping and enrichment analyses with permutation testing. To enhance the interpretability of our gene set analysis results, we clustered significant GO terms with nominal p-value threshold ≤ 0.05 based on their semantic similarity using rrvgo R package^45^(**see Supplementary Materials**).

### Ethics statement

Ethical approvals were obtained from UKB (11/NW/0382) and CLSA (190213), with institutional approvals from McGill University (A03-M20-21B, A05-M25-19A).

## Results

### Cross-sectional analysis of chronic pain and asthma traits

Supplementary Figure 1 presents a multi-step analytical pipeline to investigate the genetic basis and biological mechanisms underlying chronic pain traits and asthma considering age-of-onset strata. Our integrative framework combines genetic association, risk and causal modeling, and functional exploration to disentangle the shared and distinct genetic mechanisms linking the two sets of traits (**Supplementary Fig. 1**). We first assessed the co-occurrence of chronic pain and asthma in 456,958 UKB participants of European descent (**Supplementary Fig. 1A-i**). We identified significant associations across 39 pairwise combinations of chronic pain and asthma phenotypes (**Fig. 1A & B**). Asthma risk was highest for late and lowest for childhood asthma. Among pain traits, widespread pain showed the highest asthma risk (3-fold increase with late asthma). For late asthma, risk increased with number of pain sites, peaking in the top five-to-seven sites stratum (*OR* = 3.02, 95% *CI* (2.65-3.46), with similar patterns for adult and childhood asthma (**Fig. 1B)**. Chronic pain risk estimates increased markedly from childhood to adult asthma and moderately from adult to late asthma, with non-overlapping 95% confidence intervals between childhood and late categories (**see Supplementary Result 1, Supplementary Table 1,2,3**).

**Figure 1.**
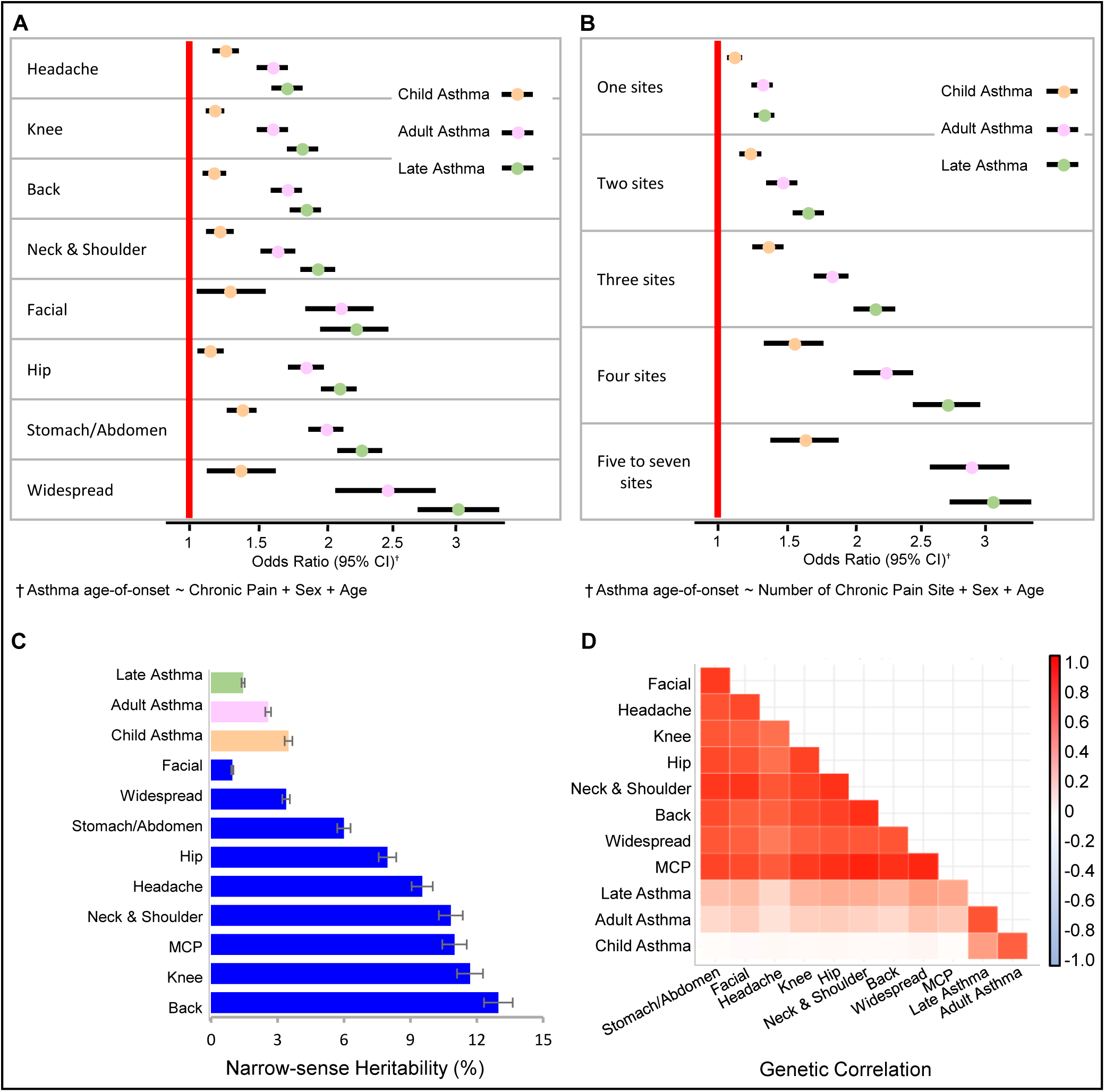
Phenotypic and genome-wide genetic parameters across asthma and chronic pain traits. **(A)** Associations between asthma stratified by age-of-onset (childhood, adult, late) and chronic pain traits, including body site-specific pain and widespread pain. **(B)** Associations between asthma age-of-onset groups and the number of chronic pain sites. Red vertical lines in panels A and B indicate no association (OR = 1). Circles represent estimated odds ratios (ORs), with horizontal black lines indicating 95% confidence intervals. **(C)** Narrow-sense heritability estimates for childhood asthma (light orange), adult asthma (pink), late asthma (light green), and chronic pain traits (blue). **(D)** Genetic correlations between asthma (stratified by age-of-onset) and chronic pain traits. The heatmap shows all pairwise correlations, with red indicating positive and blue indicating negative correlations; darker hues represent stronger effects.

### Genome-wide association scans across chronic pain and asthma traits

To characterize the shared and distinct genetic architecture across the nine chronic pain phenotypes and three asthma phenotypes, we first performed genome-wide association analyses, and identified 179 distinct loci (**Supplementary Fig. 1A-ii, Supplementary Table 4**). Five loci overlapped between at least one chronic pain and one asthma phenotype. All five included childhood asthma, one included adult asthma and headache (with nearest gene to lead SNP *SDR9C7-TMEM194A*), three included headache, one included knee and back pain (*HSD17B8-IP6K3*) (**see Supplementary Result 2, Supplementary Fig. 2**).

### Heritability, genetic correlation, and genetic architecture of chronic pain and asthma traits

For chronic pain, SNP-heritability ranged from 2.4% for facial pain to 11% for multisite chronic pain (MCP), a quantitative trait counting the number of body sites with a report of chronic pain, with most musculoskeletal pain traits showing heritability at 6-8.5%. For asthma, heritability decreased with age-of-onset: 3.5% for childhood, 2.6% for adult and 1.5% for late asthma (**Supplementary Fig. 1B-i**, **Fig. 1C, Supplementary Table 5**). Pain traits showed high genetic correlations with MCP (*rg* = 0.81-0.96), demonstrating MCP’s effectiveness at capturing features of the genetic architecture of individual pain sites (**Fig. 1D, Supplementary Table 6**). Adult asthma was more strongly genetically correlated with both late and childhood asthma (*rg* = 0.78 and 0.83 respectively) than childhood and late asthma (*rg* = 0.50). Pain traits showed the strongest genetic correlation with late asthma, particularly with widespread pain (*rg* = 0.50) and MCP (*rg* = 0.45), with correlations decreasing from late to childhood asthma.

We investigated the genetic architecture of chronic pain and asthma traits to estimate polygenicity—the number of causal variants contributing to trait heritability—and discoverability—the average strength of those variant effects—under univariate modeling assumptions (**Supplementary Fig. 1B-ii-iii**). Chronic pain traits exhibited high polygenicity and low discoverability, an inverse trend compared to asthma traits (**Fig. 2A & B, Supplementary Table 7**). Late-onset asthma resembled chronic pain in its polygenicity-to-discoverability ratio, Late-onset asthma showed a similar polygenicity-to-discoverability ratio as chronic pain, while that for childhood asthma reflected larger-effect variants.

**Figure 2.**
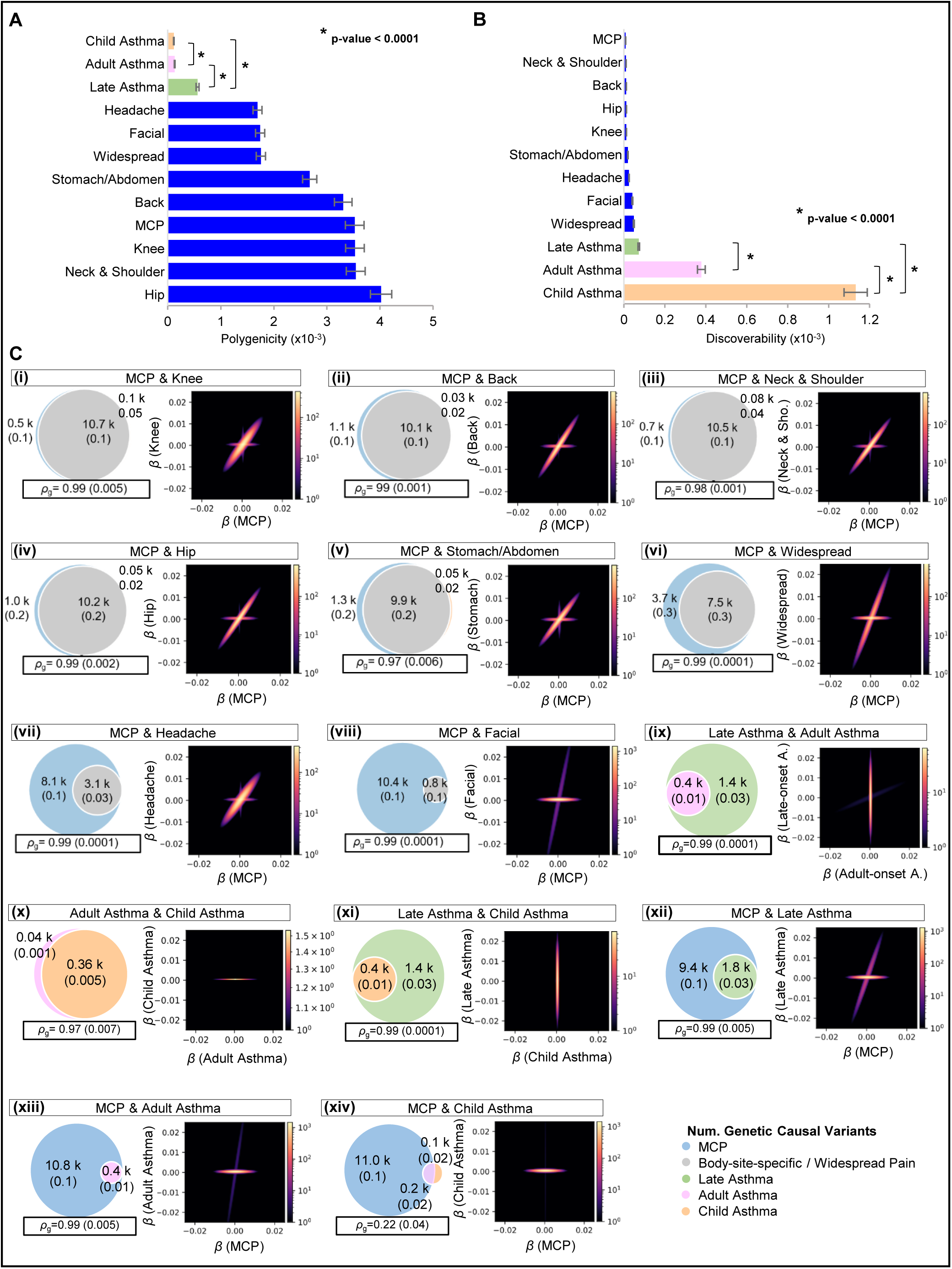
Common polygenic architecture underlying chronic pain and asthma traits. **(A)** Estimated polygenicity across chronic pain and asthma traits. **(B)** Discoverability estimates, reflecting average effect sizes among causal variants. **(C)** Shared genetic architecture illustrated by Venn diagrams and heatmaps. Venn diagrams show the number of unique and overlapping causal variants between pairs of traits. Heatmaps display bivariate density of allele substitution effect sizes (beta) for each SNP pair, with color intensity scaled by log (N), where N is the number of SNPs per bin. Panels (**i–viii**) show comparisons between MCP and site-specific or widespread chronic pain traits; (**ix–xi**) show comparisons across asthma subtypes; and (**xii–xiv**) compare MCP and asthma traits.

The number of estimated causal variants ranged from 0.8 to 11.2*K* for chronic pain traits. Most musculoskeletal pain and headache variants were contained within the MCP set, highlighting MCP’s broad genetic representation (**Fig. 2C i-viii**). Childhood and adult asthma shared 90% of causal variants. Late asthma included all those variants plus 1.4*K* trait-specific ones (over 70%) (**Fig. 2C ix-xi**). Moreover, all causal variants for late asthma (1.8*K*) were nested within MCP’s larger set, while childhood asthma (0.4*K*) shared only 66% of its variants with MCP (**Fig. 2C xii-xiv**) under bivariate modeling assumptions.

MCP-late asthma comparisons showed highly correlated observed and model-estimated GWA scan effect sizes, indicating shared genetic architecture, while MCP-childhood asthma comparisons showed no correlation. We confirmed the goodness of fit of the MiXeR model to our data in univariate and bivariate analyses (**Supplementary Table 8**).

### Meta-analyses across chronic pain and asthma trait pairs

We conducted pairwise meta-analyses using MTAG, enhancing statistical power for each chronic pain or asthma trait by conditioning on a trait from the other category. The conditional effect was most notable for late asthma when boosted by MCP, as measured by the difference in mean χ2 values between the boosted and straight GWA scans (Delta χ2), while the magnitude was lowest for childhood asthma (**Supplementary Fig. 3A-C**). Mean χ² values were higher for MCP compared to other chronic pain traits. These values did not substantially increase through the inclusion of a secondary asthma trait, although late asthma showed a slight boosting effect on MCP (**Supplementary Fig. 3D-F**).

### Cross-trait polygenic risk score predictions

After validating UK Biobank PRS in Canadian Longitudinal Study on Aging (CLSA) samples for chronic pain and asthma phenotypes (**Supplementary Fig. 1B-iv, Fig. 3A & B, Supplementary Table 9**), we tested whether MCP-based polygenic risk scores could predict asthma. In the UKB, asthma risk increased from the 60th percentile of MCP PRS onward, with the strongest effects for late asthma and the weakest for childhood asthma (**Fig. 3C**). The CLSA showed a similar pattern: late asthma showed significant prediction above the 80th percentile, adult asthma above the 90th percentile, and childhood asthma showed no association (**Fig. 3D**). In the reverse direction, asthma PRS did not predict chronic pain traits, except for knee pain in the CLSA, where late asthma PRS was associated with increased risk above the 80th percentile (**Supplementary Fig. 4A–C**).

**Figure 3.**
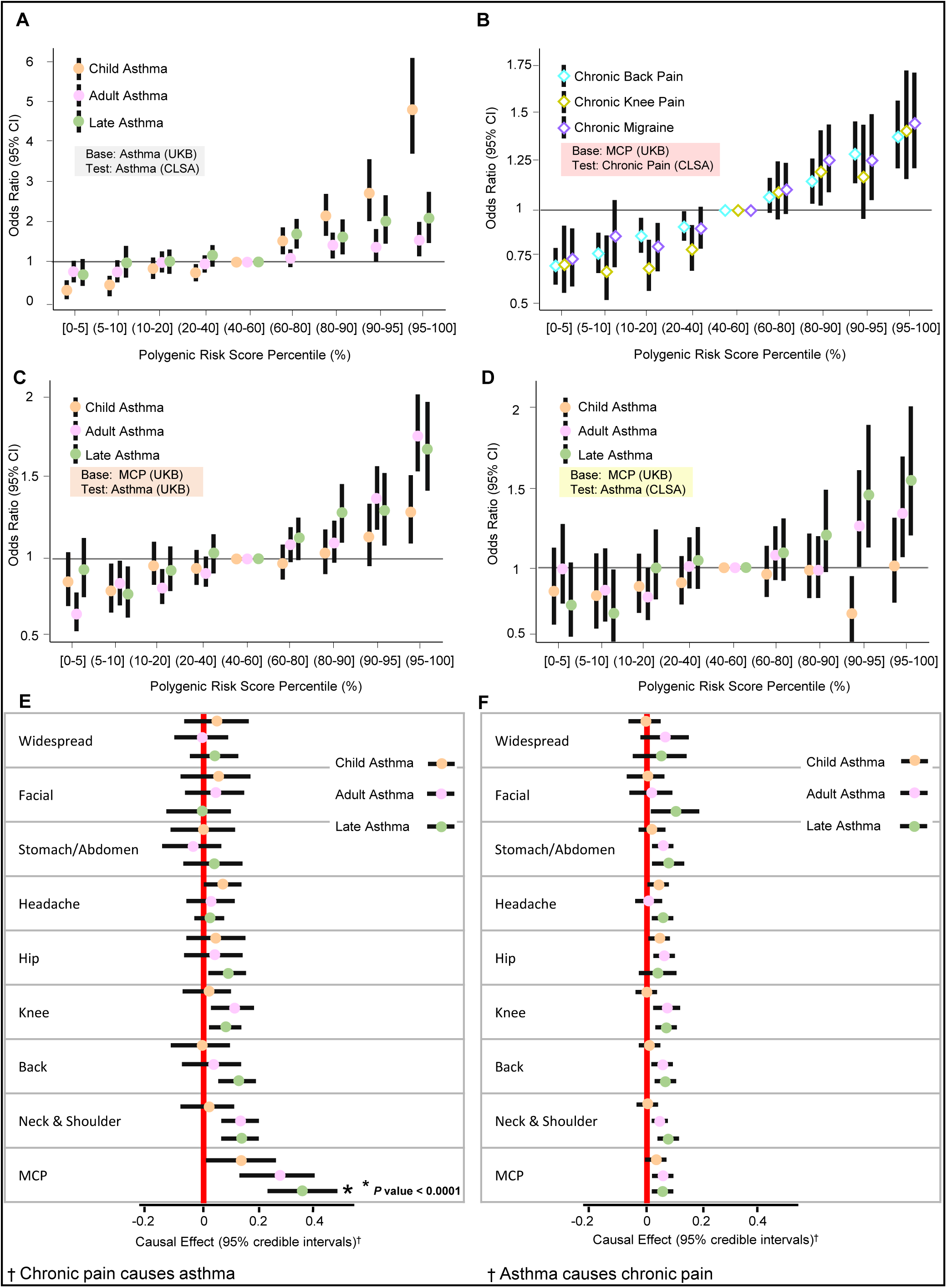
Predictive power of polygenic risk scores (PRS) and bidirectional causal modeling between chronic pain and asthma strata. (A–D) Risk of asthma or chronic pain across PRS percentile bins (<5% to >95%) in the UK Biobank (UKB) and Canadian Longitudinal Study on Aging (CLSA). Horizontal black lines indicate no association (OR = 1); vertical black lines show 95% confidence intervals. **(A)** Risk of asthma strata in the CLSA across asthma PRS percentiles. **(B)** Risk of chronic migraine, back pain, and knee pain in the CLSA across MCP PRS percentiles. **(C)** Risk of asthma strata in the UKB across MCP PRS percentiles. **(D)** Risk of asthma strata in the CLSA across MCP PRS percentiles. **(E–F)** Bidirectional Mendelian randomization (MR) using CAUSE. **(E)** Causal effects of chronic pain traits on asthma strata. **(F)** Causal effects of asthma strata on chronic pain traits. Vertical red lines indicate no causal effect (γ = 0); circles mark γ estimates with horizontal black lines showing 95% confidence intervals.

### Causal direction using mendelian randomization and longitudinal analyses

We used bidirectional Mendelian randomization (CAUSE) to assess causal and pleiotropic effects between chronic pain and asthma traits **(Supplementary Fig. 1C-i)**. After Bonferroni correction, only MCP showed a significant causal effect on late asthma (γ = 0.35, *P-Bonf.* = 0.032)(**Fig. 3E, Supplementary Tables 10, 11**). No significant causal effects were found in the reverse direction (**Fig. 3F**). This directional relationship was supported by converging evidence from longitudinal analyses in the UKB and CLSA cohorts, reinforcing the MR finding that chronic pain precedes and contributes to late-onset asthma (**Supplementary Fig.01 C-ii-iii**). Among individuals initially free of asthma, baseline chronic pain status predicted late asthma development across all body sites, (relative risk (*RR*): 1.67-3.35) except for chronic widespread pain (*RR* = 2.12, *CI*: 0.77-5.86) (**Fig. 4A, Supplementary Table 12**). This causal relationship was further supported by increasing risk observed with increasing numbers of pain sites at baseline (*RR*:1.57-3.35) (**Fig. 4B**). In contrast, baseline late asthma showed weaker associations with subsequent chronic pain development, with the highest risks observed for neck/shoulder pain (*RR*=1.53, *CI*:1.07-2.19) and hip pain (*RR*=1.83, *CI*:1.19-2.82) in the UKB, and for migraine (*RR*=1.91, *CI*:1.40-2.60) in the CLSA (**Fig. 4C**). Baseline asthma did not predict the number of pain sites at follow-up (**Fig. 4D**), suggesting asymmetry in the relationship.

**Figure 4.**
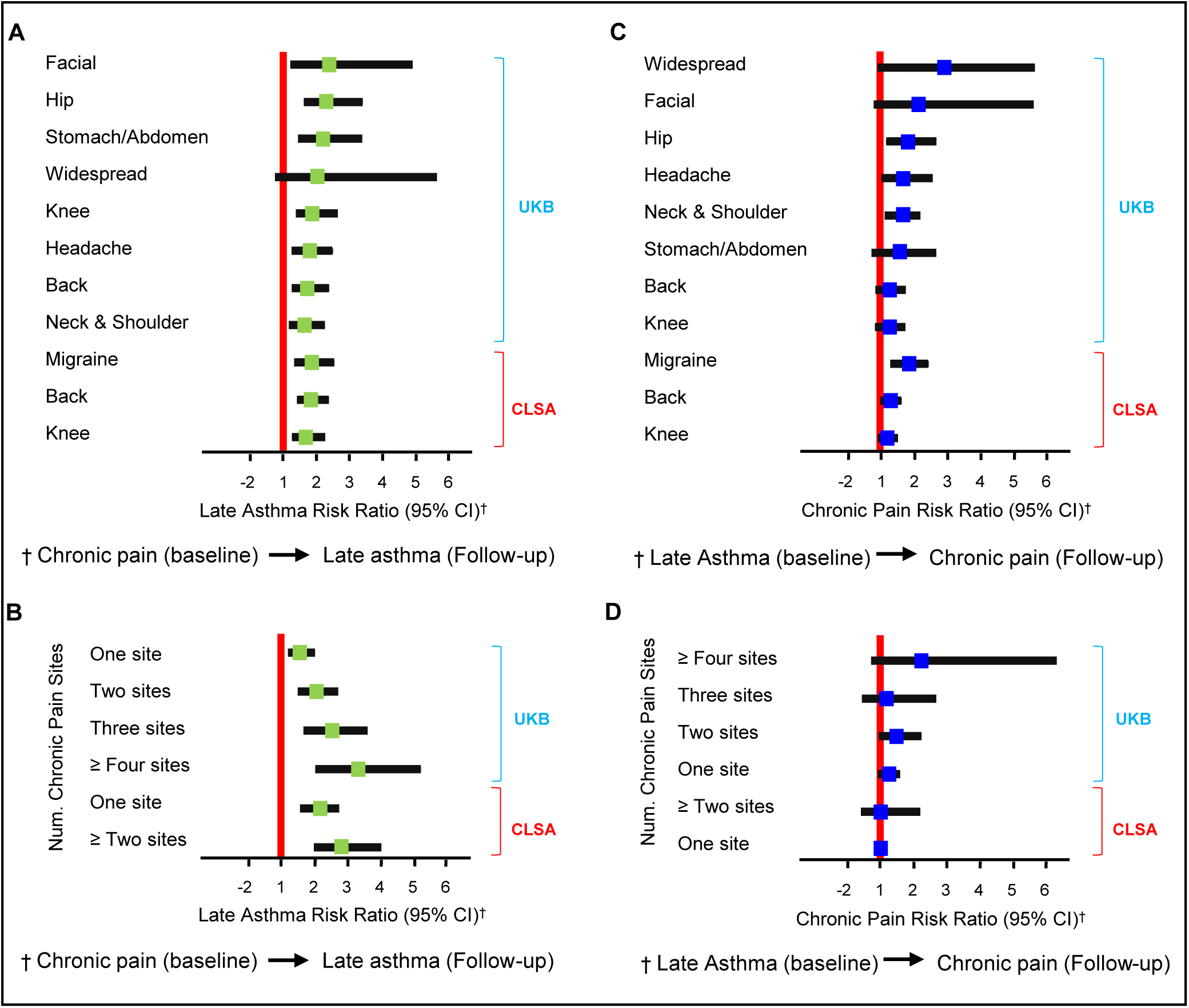
Longitudinal analyses for late asthma and chronic pain traits. **(A)** Risk of late asthma at follow-up among individuals with vs. without chronic pain at baseline. **(B)** Dose-response relationship between the number of chronic pain sites at baseline and risk of late asthma at follow-up. **(C)** Risk of chronic pain at follow-up among individuals with vs. without late asthma at baseline. **(D)** Dose-response relationship between baseline late asthma and risk of chronic pain at follow-up. Vertical red lines indicate no risk (RR = 1); horizontal black lines represent 95% confidence intervals.

### Biological function

To biologically interpret the causal relationship between MCP and late asthma, we performed pathway, tissue, and cell type enrichment analyses (**Supplementary Fig. 1D-i-ii**). Gene-based tests (**Fig. 5A**) revealed that MCP boosting increased the number of significant asthma genes. For late asthma, 41.4% (141 genes) were significant (*FDR* ≤ 0.1), compared to only 7.7% (29 genes) for adult and 3.5% (39 genes) for childhood asthma, attributable to boosting by MCP (**Fig. 5B i-iii, Supplementary Table 13**).

**Figure 5.**
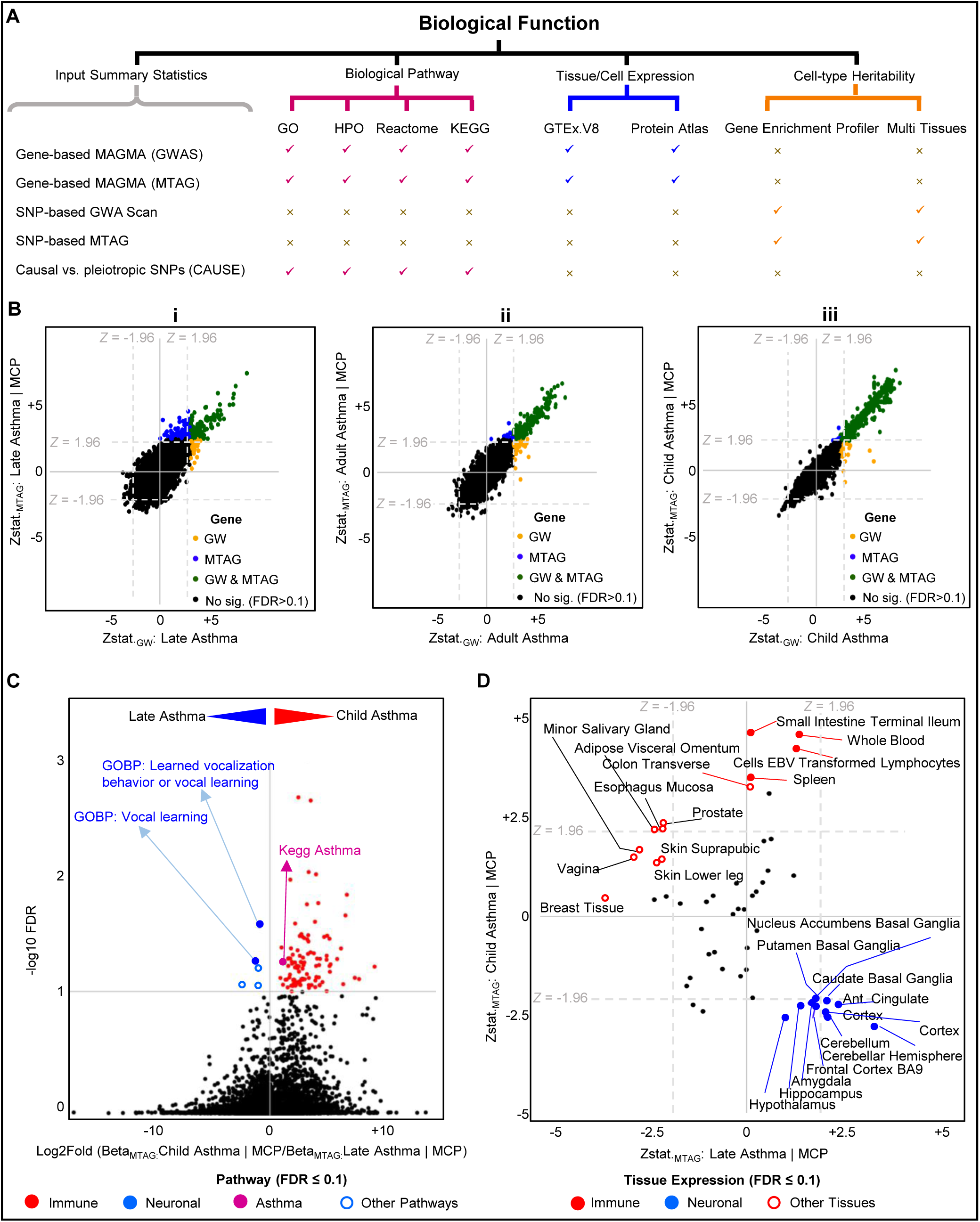
Functional characterization of shared genetic architecture between chronic pain and asthma. **(A)** Input files included gene and SNP lists from trait-specific GWA scans and MTAG summary statistics, gene-based MAGMA results, and SNP effect estimates from CAUSE. Pathway analyses were performed using Human Phenotype Ontology (HPO), Reactome, Gene Ontology (GO), and KEGG. **(B)** MAGMA gene-based results comparing trait-specific GWA scans with MTAG summary statistics of asthma age-of-onset strata boosted by MCP. Quadrant plots display Z-statistics, with dashed vertical and horizontal lines indicating nominal significance thresholds. Panels: **(i)** late asthma, **(ii)** adult asthma, **(iii)** childhood asthma. **(B)** MAGMA gene-set enrichment analysis using MTAG results for childhood and late asthma, both boosted by MCP. The volcano plot shows log fold change in gene-set effect sizes (x-axis) versus –log FDR-adjusted p-values (y-axis); the horizontal line marks the FDR threshold. **(D)** MAGMA gene-property analysis of tissue enrichment using GTEx v8 and MTAG summary statistics for childhood and late asthma (boosted by MCP). Quadrant plot Z-statistics are shown, with dashed lines marking nominal significance thresholds.

Pathway enrichment analyses of GWA summary statistics identified 43 MCP-associated pathways (mostly neurological) and 27 shared asthma pathways (*FDR* ≤ 0.1)(**Supplementary Table 14**). Shared asthma pathways were primarily immune-related, including interleukin-1 receptor activity, the KEGG asthma pathway, and MHC class II function. Notably, the only nervous system pathway—learned vocalization behavior—was shared between MCP and late asthma.

To identify functional differences by asthma age-of-onset, we compared pathway enrichment beta values between MCP-boosted late and childhood asthma. Among 113 significantly enriched pathways, 99 were immune-related and more strongly enriched in childhood asthma. In contrast, late asthma showed enrichment for two nervous system pathways—vocalization behavior and vocal learning—and three additional pathways (**Fig. 5C, Supplementary Table 15**).

We next used tag SNPs from CAUSE (delta ELPD < 0, *N*=2,743) to identify pathways enriched for causal effects of MCP on late asthma (**Supplementary Tables 16, 17**). Gene Ontology and Human Phenotype Ontology analyses revealed 18 (4 parent-node level clusters) and 6 significant pathways, respectively, all involving nervous system or musculoskeletal function (**Supplementary Fig. 5A, Supplementary Tables 18, 19**). The top 12 causal genes (delta ELPD ≥ –0.08), including *DCC, GMPPB-AMIGO3-RNF123*, and *FOXP2*, were consistently represented across the enriched neurological pathways. Several of these genes were also highly boosted by MCP in the meta-analysis (**Supplementary Fig. 5B**). In contrast, no significant pathways were identified using pleiotropic variants (delta ELPD > 0, *N*=1,972).

Tissue enrichment analyses using GTEx highlighted 13 nervous system tissues—including cerebellum, frontal cortex, and basal ganglia—for both late asthma and MCP, while childhood and adult asthma were enriched in immune tissues such as whole blood, EBV-transformed lymphocytes, and small intestine (**Supplementary Tables 20–21**). Comparing late and childhood asthma boosted by MCP, immune tissues were highly enriched in childhood asthma, while brain regions were significantly enriched for late asthma (*FDR* ≤ 0.1)(**Fig. 5D, Supplementary Table 21**). MCP-boosted late asthma additionally showed enrichment in cerebellar substructures—cortex, vermis, and flocculonodular lobe—based on single-cell RNA data (Human Protein Atlas) (**Fig. 6A-i–ii**), while other asthma strata showed no such enrichment (**Fig. 6A-iii–vi**). Cell types enriched in late asthma through MCP boosting included microglia, astrocytes, excitatory and inhibitory neurons, and oligodendrocytes, with only microglia also enriched in MCP-boosted adult asthma (**Supplementary Table 22**).

**Figure 6.**
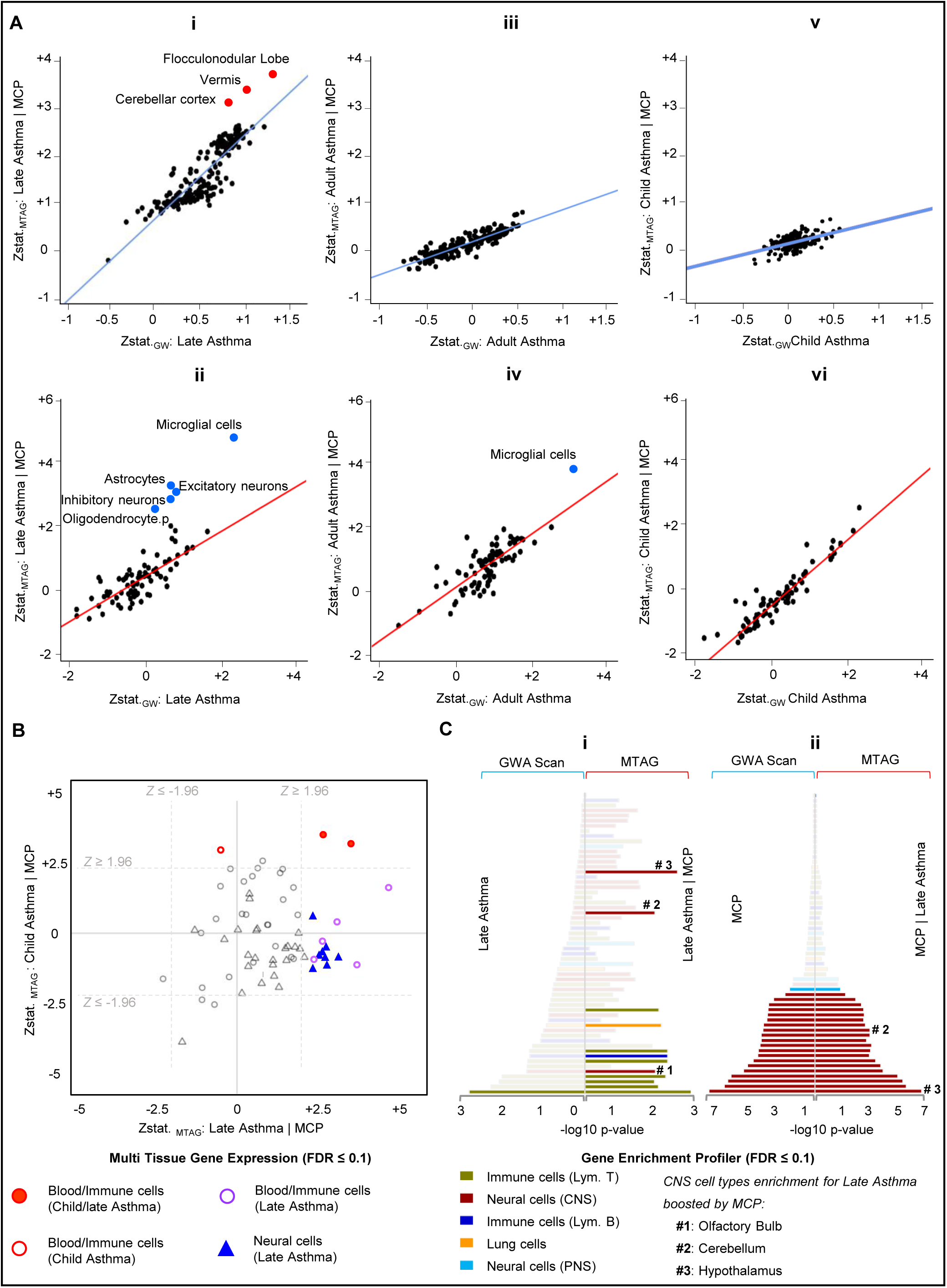
Differential enrichment of immune and central nervous system signals in childhood and late asthma. **(A)** Brain tissue and cell-type enrichment using data from the Human Protein Atlas and both trait specific GWA scans and MTAG summary statistics of asthma age-of-onset strata boosted by MCP. Panels show results for asthma traits. **(i–ii)** Late asthma. **(iii–iv)** Adult asthma. **(v–vi)** Childhood asthma. **(B)** Cell-type-specific partitioned heritability enrichment based on multi-tissue gene expression data using MTAG summary statistics for childhood and late asthma (both boosted by MCP). Quadrant plots show Z-statistics, with dashed vertical and horizontal lines indicating nominal significance thresholds. **(C)** Cell-type-specific heritability enrichment using the Gene Enrichment Profiler dataset. **(i)** Pyramid plot shows –log p-values for late asthma (boosted by MCP), with GWA scan results on the left and MTAG results on the right. **(ii)** Pyramid plot shows –log p-values for MCP (boosted by late asthma), with GWA scan results on the left and MTAG results on the right.

Cell-type heritability analysis using Multiple-tissue Analysis of Gene Expression revealed enrichment for both late and childhood asthma in broad lymphocytes and T lymphocytes (*FDR* ≤ 0.10)(**Fig. 6B**, solid red dots), with lymphocyte-null cell types enriched only in childhood asthma (red ring). Late asthma showed unique enrichment in monocytes, phagocytes, the mononuclear-phagocyte system, and bone marrow cells (**Fig. 6B**, purple rings), as well as in CNS tissues, namely cerebellum, metencephalon, cerebellar hemisphere, limbic system, and brain stem cells (**Fig. 6B**, blue triangles, **Supplementary Table 23)**.

Gene Enrichment Profiler confirmed heritability enrichment for MCP-boosted late asthma in T and B lymphocytes, CNS regions (cerebellum, hypothalamus, olfactory bulb), and respiratory tissues (**Fig. 6C-i**). In contrast, MCP alone showed enrichment in 20 CNS tissues, but no additional enrichment when boosted by late asthma, reinforcing the asymmetry in causal direction (**Fig. 6C-ii, Supplementary Table 24**).

## Discussion

We provide the first comprehensive genetic dissection of the link between chronic pain and asthma, revealing a complex, age-dependent relationship driven primarily by shared architecture between MCP and late-onset asthma. Multiple lines of evidence—including polygenic risk scores (PRS) in two large cohorts (UK Biobank and CLSA), longitudinal prediction of asthma onset by baseline chronic pain, and Mendelian Randomization (MR)—consistently supported a directional causal effect from MCP to late asthma. The substantial boosting effect through meta-analysis of late asthma by MCP highlights a shared polygenic component, characterized by increased late asthma effect sizes across the genome, as supported by bivariate causal mixture models. Pathway and cell-type heritability analyses indicated immunological function for childhood asthma, while revealing both immunological and substantial neurological contributions for late asthma. Together, these findings suggest a biologically coherent and directionally causal relationship between chronic pain and late asthma, underpinned by shared neurological and immune mechanisms that emerge in mid-to-late life.

This work expands upon previous reports of a more modest genetic correlation (∼20%) between MCP and asthma as a broad phenotype^17,24,25^ by demonstrating a substantially higher genetic correlation between MCP and late asthma (*rg* = 0.45), with correlations decreasing progressively from late to childhood asthma. These findings align with an earlier study showing genetic correlations between adult asthma and insomnia or depressive symptoms—traits that commonly co-occur with chronic pain—while no such correlation was observed for childhood asthma^46^. While a prior MR study suggested a horizontally pleiotropic relationship between MCP and asthma as a broad phenotype^25^, our analyses provide converging evidence for a directional causal effect of chronic pain on late asthma (γ = 0.35).

Previous UK Biobank GWA studies have independently examined asthma stratified by age-of-onset or MCP for chronic pain, but this is the first study to jointly analyze these traits while incorporating internal heterogeneity on both sides. One prior study stratified asthma by the same age cut-offs we used like a recent epidemiological survey^9^. Their study found strong differences in comorbidities, asthma control, and demographic factors across these strata, reinforcing the clinical and epidemiological relevance of our categorization. Earlier studies concluded that childhood asthma has a distinct genetic profile, while adult asthma shares many of these signals but adds little unique signal of its own^46^. Our findings extend this model by showing that late asthma not only shares genetic factors with earlier-onset forms but also carries additional trait-specific variation.

Consistent with prior GWAS identifying lymphocyte-driven immune involvement in asthma^20,47,48^, we observed strong enrichment for lymphocyte-specific genetic signal in both childhood and late asthma when boosted by MCP, suggesting a shared immune polygenic component. Childhood asthma showed stronger enrichment across immune pathways and tissues, aligning with previous findings of its genetically driven, allergy-related architecture^19,49^. In contrast, late asthma genetic signals uniquely demonstrated enrichment in nervous system pathways and CNS tissues including the cerebellum, alongside broader leukocyte categories such as phagocytes and monocytes. This combined neurological and immune involvement has not been prominent in prior asthma GWA studies and highlights central nervous system involvement as a novel component of late asthma and its overlap with MCP^50^.

These findings are consistent with a dose-response pattern in genetic architecture across asthma strata, and with a biologically grounded dimorphism: functional analyses revealed exclusively immunological pathways for childhood asthma and both immunological and neurological pathways for late asthma, while no clear enrichment emerged for intermediate adult asthma. The lack of pathway enrichment for intermediate-onset adult asthma may reflect a dilution effect, combining two distinct variant sets—some shared with childhood asthma, others with late asthma—without convergent biological evidence. In this context, the shared genetic risk may represent “poor aging” variation, consistent with the increasing prevalence and frequent co-occurrence of asthma and chronic pain in later life^51,52^. More speculatively, our findings may reflect a common inflammaging-related latent phenotype—an underlying low-grade inflammatory state that manifests differently across individuals as either chronic pain, late asthma, or both^51,52^.

Our study has some limitations. First, as analyses focused on European ancestry participants, results may not generalize to other populations. Second, UK Biobank’s non-specific self-reported chronic pain classification fails to distinguish between nociceptive, neuropathic, and nociplastic mechanisms, constraining our causal interpretations, particularly across different body sites.^53^ This heterogeneity in pain mechanisms may explain the weak or non-causal effects observed for some of the body-site-specific pain traits in our longitudinal analysis. Despite these limitations, the multi-cohort design, integration of longitudinal and genetic data, and convergence across analytic approaches lend robustness to our findings.

In summary, our comprehensive analyses reveal a complex relationship between chronic pain and asthma that varies substantially by phenotypic subtype, with the strongest and most causally relevant link between MCP and late asthma. The complete overlap of causal variants, consistent directional evidence, and shared neurological pathways suggest that chronic pain may contribute to late asthma development via central neurological mechanisms, potentially involving neuroinflammatory processes that bridge cerebellar and immune systems. Our results may offer promising targets for interventions aimed not only at symptom management but at the prevention of both chronic pain and late-onset asthma.

## Authors’ contributions

A.V.G. and L.D. conceived the study. G.K. conceived and designed the analysis plan. G.K. performed GWA scans and bioinformatics analyses and generated results displays. A.V.G. and G.K. interpreted the results. A.V.G. and G.K. drafted and revised the manuscript. M.P. provided input and advice on statistical analysis using eQTL data and helped in creating a mathematical formula for causal pathway analysis. N.D, C.C., and Y.C. participated in UKB data preparation and processing. N.D., M.Z., L.H, P.H., and S.B participated in the CLSA phenotype classification and QC of CLSA genotyping data. L.D. advised on the analysis plan, interpretation of results at all stages and participated in funding acquisition. A.V.G. oversaw the analysis plan and contributed to funding acquisition. All authors critically reviewed and approved the final version of the manuscript.

## Acknowledgements

The research team would like to express their appreciation to all participants of UKB and the CLSA. Our work was made possible thanks to generous support from The Louise and Alan Edwards Foundation c/o, The Jewish Community Foundation Montreal, the Canadian Excellence Research Chair for Human Pain Genetics (CERC09), and NIH grant U54 DA049110 (held by LD). G.K. was supported by the Catherine Bushnell Pain Research Postdoctoral Fellowship, the International Association for the Study of Pain, and the Quebec Network of Junior Pain Investigators.

## Data availability

The summary statistics of all GWAS used in this study will be publicly available from GWAS Catalog.

## Code availability

https://github.com/ermia1313/Causal-Pathway/

## Resource

REGENIE (v3.1.3): https://rgcgithub.github.io/regenie/

LDSC (v1.0.1): https://github.com/bulik/ldsc

FUMA: https://fuma.ctglab.nl

PLINK: https://www.cog-genomics.org/plink/

LDLINK: https://ldlink.nci.nih.gov/?tab=home

MTAG: https://github.com/JonJala/mtag/

CAUSE: https://jean997.github.io/cause/

MAGMA: https://cloufield.github.io/GWASTutorial/09_Gene_based_analysis/

PRSice2: https://choishingwan.github.io/PRSice/

ct-LDSC: https://github.com/bulik/ldsc/wiki/Partitioned-Heritability/

MiXer: GitHub - precimed/mixer: Causal Mixture Model for GWAS summary statistics

CLSA: https://www.clsa-elcv.ca/data-collection

Gene Ontology (GO): https://www.geneontology.org/

rrvgo: http://revigo.irb.hr/

REACTOME: https://reactome.org/PathwayBrowser/

Human Phenotype Ontology (HPO): https://hpo.jax.org/

KEGG: https://www.genome.jp/kegg/pathway.html

