## Supplementary material for "Causal evidence linking chronic pain genetics to late-onset asthma via the nervous system": Online Supplemet: ONLINE_SUPPLEMENT_BJA.docx

**Author affiliations:**

**Supplementary Method:**

**Chronic Pain, Asthma, and COPD Data in the UKB**

We drew on health questionnaires and genome-wide genotyping datasets from the UK Biobank (UKB) participants (https://biobank.ndph.ox.ac.uk/showcase, last accessed Dec 2022). We classified chronic pain cases and controls in the UKB based on two consecutive questions (Resource: 113241). The first question was, "In the last month, have you experienced any of the following that interfered with your usual activities?" (data field 6159). The options provided were: headache, facial pain, neck or shoulder pain, back pain, stomach or abdominal pain, hip pain, knee pain, pain all over the body (widespread), and none of the above. The participants who answered "none of the above" to data field 6159 were classified as healthy controls. The participants who chose one or more pain sites were prompted to answer a second question per site on whether this pain lasted for more than 3 months (data fields 3799: headaches; 4067: facial pain, 3404: neck/shoulder pain; 3571: back pain; 3741: stomach/abdominal pain; 3414: hip pain; 3773: knee pain; 2956: pain all over the body). The participants who answered yes to the second question for one or more pain sites were defined as chronic pain cases. The participants who answered yes at field 2956: Pain all over the body were attributed a value of eight, which is one unit more than the seven other body sites to choose from.

Asthma status classification was drawn from data field 20002. Based on a touchscreen questionnaire, study subjects were asked whether they had been told by a doctor that they had one or more illnesses from a list including asthma (data code 1111). Next, an interviewer who had received answers from the touchscreen questionnaire asked, "In the touch screen, you selected that you have been told by a doctor that you have other serious illnesses or disabilities. Could you now tell me what they are?" Finally, the interviewer recorded the date of the first diagnosis, entering an estimated date rather than an unknown if the participant could only provide an approximate answer registered in field 20009.

We extracted doctor-diagnosed Chronic Obstructive Pulmonary Disease (COPD) from self-reported non-cancer illnesses to figure out any potential misclassifications of asthma cases that might affect the epidemiological association between asthma and chronic pain. Similar to the asthma case definition, the COPD status classification was drawn from data code 1112.

**Identification of Europeans in the UKB**

We merged genomic data from UK Biobank (UKB) participants (N = 502,599) with data from the 1000 Genomes (1KG) Project using 78,709 SNPs shared across datasets, filtered for MAF > 0.01, genotype missingness < 0.1, and LD pruning (r² < 0.15). Principal component analysis (PCA) was performed, and the first three eigenvectors accounted for the majority of variance. We modeled this 3D space as a multivariate normal distribution and used maximum likelihood estimation to define the mean and variance-covariance matrix based on 1KG European reference individuals. A threshold value was derived from this distribution, and UKB participants were classified as European ancestry if they self-identified as white and their projected PC values exceeded the threshold. We excluded individuals with mismatched genetically determined versus self-reported sex (N=355) and participants who opted out of the study, yielding a final analytic sample of 456,958 individuals. The procedure was implemented using in-house R scripts and follows the general framework of PCA-based ancestry inference.

**Phenotypic associations in the UKB**

We selected random samples of cases (80%) and controls (90%) for each phenotype, including stratified asthma age-of-onset, MCP, and body site-specific chronic pain traits, separately. Then, we performed multivariate analysis using a generalized linear model (GLM) in R to test for association between chronic pain traits and asthma age-of-onset, adjusting for sex and age at recruitment. We also tested for association between asthma age-of-onset, COPD age-of-onset, and chronic pain traits. Further, we used multivariate regression analysis to test for any association between chronic pain and asthma traits in the absence of COPD, adjusting for sex and age at recruitment.

**Genome-wide association scans in the UKB**

Genome-wide association (GWA) scans were performed for each trait—random samples of cases (80%) and controls (90%)— at baseline by fitting linear mixed models in two steps as implemented in Regenie (v3.1.3) . In the first step, the whole genome regression model was fitted in 200 blocks of 577,232 genotype markers subsets to reduce data dimensions. We applied ridge regressions on MCP (linear), body site-specific chronic pain, and asthma age-of-onset (logistic) to obtain a reduced model with 2,886 predictors for each trait. Next, a second level of relevant linear or logistic ridge regression analysis was carried out for a range of shrinkage parameters using the method of stacked regression to control for over-fitting due to the positive correlation between each predictor and the phenotype. Then, the best genetic predictor value was estimated through combining predictors by applying cross validation scheme. Finally, we constructed genetic prediction on 22 autosomal chromosomes by leave-one-chromosome-out (LOCO) approach. In the second step, the effect of each imputed variant was tested using LOCO predictions as a covariate by corresponding regression analysis (logistic/linear) and making a balance between case and control samples via saddle-point approximation. To account for multiple tests, the standard genome-wide threshold of 5 x 10^−8^ was used to identify a significant association between a genetic variant and a trait.

**Lead SNPs identification in the UKB**

Trait-specific summary statistics of the GWA scan were analyzed using the online platform for functional mapping and annotation of GWA studies (FUMA v.1.4.1) to annotate independent signals and corresponding risk loci. First, significant independent SNPs with r^2^ < 0.6 surpassing the genome-wide threshold (p-value) were identified. For each independent significant SNP, all candidate SNPs in high pairwise LD (r^2^ > 0.6) present either in the input data or in the reference panel (European sample of 1KG phase 3), were included to define a genomic risk locus. Next, a strict threshold at r^2^ < 0.1 was applied to define independent lead signal(s), and the signal with the lowest p-value was defined as the top lead SNP among independent significant SNPs. Then, all SNPs in high pairwise LD (0.1 > r^2^ < 0.6) were assigned to the same LD block. The contiguous blocks were merged into one independent locus if the distance between them was less than 250 kb, otherwise blocks were different loci. We implemented a similar process of gene prioritization on annotated results using FUMA. The gene mapping was not limited to lead variants, so we applied functional consequences of all candidate SNPs in LD by ANNOVAR to assign positionally nearest protein-coding genes at a maximum distance of 10 kb, which were obtained from Ensemble build v92.

**Heritability, genetic correlation, and trait-specific and cross-trait genetic architecture**

Single-trait SNP-based heritability and cross-trait genetic covariance and correlation between chronic pain and asthma phenotypes were estimated using LDSC. We employed 1KG phase 3 (European) as the reference panel population and used the subset of non-HLA HapMap3 SNPs (1.2M). We quantified the genetic architecture components of chronic pain and asthma age-of-onset to determine the number of unique and overlapping causal variants for pairs of traits using MiXeR. MiXer fits genetic architecture parameters, including effective causal strength of association (discoverability) and the proportion of causally associated SNPs (polygenicity). The parameters are optimized to reflect a best-observed set of Z statistics from the original GWA scan summary statistics for each trait by constructing the model on HapMap3 SNPs and testing this on the full set of markers. For each trait, HapMap3 SNPs with MAF > 0.05, imputation Info score > 0.3 were used (N=1,169,850) after excluding ambiguous SNPs, Major Histocompatibility Complex (MHC) on chromosome 6 (6:26000000-34000000), and other long-range LD regions. Then, the estimated parameters were tested on 11,015,833 genetic variants with MAF > 0.002, incorporating detailed LD structure information from the 1KG phase 3 dataset on 489 subjects with European ancestry (reference panel). The mean and standard deviation of estimated parameters from the fitted and testing models are extracted separately after iterating each model 20 times. Next, the number of trait-specific causal variants (*Nc*) is estimated by multiplying Polygenicity (*π*) by the number of SNPs in the reference panel in the testing model.

*Nc* = *π* × *Num.* SNPs

Extending the single trait causal mixture model to the bivariate model to quantify polygenic overlap (two traits)^57^, MiXeR is built on four bivariate normal distributions: Two distributions model causal components for variants unique to each trait, and another model causal components for variants shared between the two traits, and the last model component for variants with no effect on either trait. Similar to single trait analysis, we used the reference panel and precomputed LD structure information from the 1KG phase 3 dataset and estimated the number of shared causal variants by polygenic overlap across chronic pain and asthma age-of-onset traits through fitting the model on HapMap3 SNPs. Next, we tested the parameters on all SNPs in the panel, iterating each model 20 times and converging the estimated parameters, separately.

**Meta-analyses in the UKB**

We conducted pairwise meta-analyses of trait-specific summary statistics of MCP, body-site-specific chronic pain, and asthma age-of-onset traits in the UKB using MTAG. MTAG implements an extended inverse-variance-weighted meta-analyses approach in the presence of overlapping samples. The method was used as it enhanced the statistical power of association test effect estimates for the first trait by integrating information from the distribution of effect size estimates and the variance of GWA scan estimates from the second trait. The MTAG-based effect sizes were adjusted for LDscore’s intercept, thereby making them robust against cryptic relatedness and population stratification. Overall, MTAG resulted in power gain shared loci relative to each trait-specific GWA scan, as it provided separate trait-specific estimates conditional on the presence of the other trait. The False Discovery Rate (*FDR*) is calculated to avoid false positive association (an inflated type I error rate) due to larger GWA scan estimates for high-powered traits. The same procedure as described above was used to define independent loci and prioritize FUMA’s protein-coding genes.

**Polygenic risk scores**

We used summary statistics of GWA scan from the UKB as the base population to construct the Polygenic Risk score (PRS) for each MCP and asthma age-of-onset phenotype. PRSice built the PRS, using the threshold and clumping approach on HapMap3 SNPs with the following 250 *kb* distance between LD blocks, clump *r^2^*>0.1, 10,000 permutations, and adjusting for age, sex, and top 40 genetic PCs. In the first round of PRS testing, we used the remaining samples (not included in the primary GWA scan) from UKB set aside as the target population, phenotype by phenotype. Next, we tested the MCP PRS on asthma age-of-onset status in the target population. We tested for an association between MCP PRS and asthma age-of-onset status, and also asthma PRS and chronic pain status using logistic regression by binning individual PRS values according to percentile ranges.

We evaluated the validity of our constructed PRS models across phenotypes based on the UKB using the CLSA cohort as the target population. A similar scheme was used, starting with the asthma age-of-onset phenotypes in the UKB to construct the PRS and testing their association with chronic pain traits in the CLSA.

**Canadian longitudinal study on aging (CLSA)**

We used the CLSA cohort as an independent population to test for PRS and replication of discovery results, including significant variants from GWA scans and longitudinal causality analyses. The CLSA is a national, ongoing study of 51,338 Canadians aged 45 to 85 years at enrolment, which started in 2010. It comprised two complementary cohorts: 1) A comprehensive cohort composed of 30,097 study subjects who were randomly selected from seven provinces, interviewed in person via home or site visits, and provided blood and urine samples; 2) A tracking cohort composed of 21,241participants who were interviewed by telephone. Data collection for the comprehensive cohort has been completed for three time points, and it will be continued until 2033. We used only baseline (2012-2015) and first follow-up data (2015-2018) for the validation of PRS and longitudinal analyses results.

**Identification of Individuals of European Ancestry in the CLSA**

A total of 26,622 non-duplicated genotyped samples from the comprehensive cohort at baseline passed quality control based on an average call rate of 95%. Imputation was performed using the TOPMed reference panel on 653,729 genotyped markers after filtering out SNPs with higher than 0.05 missing rate, MAF of ≤0.0001, discordant genotype frequency between batches, departure from HWE, discordance of genotyping across control replicates, sex genotype frequency discordance, and unmatched alleles in the human genome GRCh37 reference sequence. We extracted 25,275 participants classified as of European ancestry using the first three genetic ancestry PCs from an analysis incorporating phase 3 of the 1KG project dataset. Because genomic positions were reported in reference to human genome build GRCh38/hg38 and lacked dbSNP’s canonical SNP identifiers, we converted the coordinates to human genome build GRCh37/hg19 and assigned dbSNP canonical identifiers using the Bioconductor package ("SNPlocs.Hsapiens.dbSNP151.GRCh38"), which yielded 8,200,520 genetic markers to perform PRS analysis.

**Chronic pain and asthma status in the CLSA**

We extracted data from the comprehensive cohort questionnaires administrated at baseline and first follow-up (FU1). The classification of chronic pain cases and controls started with the back pain question, "Have you ever had pain in your back on most days for at least one month?" (baseline data field "OST-BP-DCS"; follow-up "OST-BP-COF1"). The participants who chose "Yes" were prompted to answer a second question on how long you had this pain (baseline data field "OST-BP-DUR-DCS"; follow-up "OST- BP-DUR-COF1"). The participants who answered "more than 3 months" at the second question at either baseline or FU1 were defined as chronic back pain cases. The participants who answered "No" to the first question at baseline and FU1 visits were classified as healthy controls. For knee pain, the question was, "During the past 4 weeks, have you had knee pain on most days?" (baseline data field "OSK-PAIN-DCS"; follow-up "OSK-PAIN-COF1"). The participants who reported persistent knee pain at both visits were defined as chronic knee pain cases. The participants who answered "No" at the baseline and FU1 were classified as healthy controls. Chronic migraine status classification was drawn from records of self-reported chronic conditions (baseline data field "CCC-MGRN-DCS"; follow-up "CCC-MGRN-COF1"). Based on the questionnaire, study subjects were asked whether they had been told by a doctor that they had migraine headaches. The participants who reported medical doctor-diagnosed migraine in two consecutive phases of the study were considered chronic migraine cases. The participants who answered "No" at both visits were classified as healthy controls. Asthma status classification was drawn from records of self-reported Chronic Airflow Obstruction (baseline data field "CCC-ASTHM-DCS"; follow-up "CCC-ASTHM-COF1"). Based on the questionnaire, study subjects were asked whether they had been told by a doctor that they had asthma (baseline data field "CCC-ASTHM-DCS"; follow-up "CCC-ASTHM-COF1"). The participants who reported medical doctor diagnosed asthma in either baseline (prevalent case) or follow-up (incident case) were considered as asthma cases. The participants who answered "No" at both visits were classified as healthy controls. We further stratified asthma cases in three age categories like the UKB (baseline data field "CCC-ASTHMAGE-DCS"; follow-up: "CCC-ASTHMAGE-COF1").

**Mendelian Randomization**

We performed MR analysis to assess whether the effects of associated signals shared between trait-specific GWA scan of chronic pain and asthma age-of-onset in the UKB were pleiotropic or mediated by one of the traits (causative) using Causal Analysis Using Summary Effect (CAUSE). This method incorporates test statistics from all GW significant and non-significant variants in the presence of overlapping samples, extracts independent variants with *r^2^*> 0.01, and lowest p-value 0.001 after pruning for LD. The effect of each genetic variant is decomposed into correlated pleiotropy (proportion of heritability mediated by confounder), uncorrelated pleiotropy (proportion of heritability explained by genetic variants directly), and causal effect (proportion of heritability mediated by another trait), and the expected log point-wise posterior density (ELPD) is tested in two nested models; sharing model (causal effect= 0) and causal model (causal effect > 0). In a test for ELPD difference (Delta ELPD), SNPs with negative and positive values are recognized as causal and pleiotropic variants, respectively. The MR analyses p-values in either sharing or causal models were corrected for multiple testing via the Bonferroni method.

**Longitudinal Analyses**

We drew on the UKB and CLSA to consider chronic pain and late asthma case status at two time points, baseline and first follow-up. We fitted logistic regression models adjusting for age and sex using R’s glm function. For each individual chronic pain and late asthma at baseline, we tested for risk of disease by status of the other phenotype at the first follow-up. For MCP, we considered a dose-response relationship between the count of chronic pain sites at baseline and the occurrence of late asthma at the first follow-up. A similar scheme was used, starting with the late asthma at baseline and the occurrence of chronic pain (count of pain sites) at the first follow-up.

**Biological function**

In gene set analyses, we consolidated the gene sets from Human Phenotype Ontology (HPO), Reactome, KEGG, and Gene Ontology (GO) terms to test any differences between the genetic association of genes inside and outside of a given gene set regarding biological pathways using gene-based p-values.

Gene property analyses were implemented to test for excess representation of specific cell types and tissues among genes associated with the trait(s) of interest with eQTL assessment across different tissue types in GTEx v8 and Human Protein Atlas datasets. We considered significant gene, tissue, and cell type enrichment after correcting for multiple comparisons (*FDR* ≤ 0.1). To assess the significance of differences in each pathway or tissue enrichment between asthma and MCP traits, we employed the following statistical approach:

$$Z=\frac{\beta_{1}- \beta_{2}}{\sqrt{SE_{1}+SE_{2}}}$$

*β1* and *β2* represent the enrichment coefficients for the first and second traits, respectively. *SE1* and *SE2* denote the standard errors associated with these enrichment coefficients. We considered significant gene-set, tissue, and cell type enrichment after correcting for multiple comparisons (*FDR* ≤ 0.1).

Human gene expression data by different cell types were obtained from Multiple-tissue Analysis of Gene Expression (72 human cell types including, central nervous system and immune and blood cell types from GTEx and Franke lab datasets), and Gene Enrichment Profiler data (Microarray data on 59 human cell types including, central/peripheral nervous systems, immune and blood systems and respiratory system). We performed cell-type specific analyses on both primary GWA scan and MTAG-boosted estimates using precomputed gene-tissue expression levels ranging from 0 to 1, of which higher expression scores (close to 1) are attributed to genes with higher expression in specific tissues. To prioritize relevant cell types, we employed 1KG phase 3 (Europeans) as the reference panel population, adjusted for LD scores, and weighted scores on the subset of non-HLA HapMap3 SNPs (1.2M) using ct-LDSC. The p-values were corrected for multiple testing (*FDR* ≤ 0.1).

We integrated the curated gene sets, GO, HPO, KEGG, and Reactome, to test any differences between the genetic association of genes inside and outside of a given gene set regarding biological pathways using CAUSE outputs. The SNPs were assigned ELPD delta values by indicating causation (negative delta) or pleiotropy (positive delta). For a given pathway, we retained the number of genes (*N*) inside the pathway that have a negative (causation test) or positive (pleiotropic test) delta, and it was tested one at a time, with causation and pleiotropy testes separately. The observed sum of values (*S-obs*) across these causative and pleiotropic genes (N) in each pathway is compared with the N genes with negative and positive delta ELPD’s values through random samplings (*S-rand*)(T = 10 million trials), respectively. The p-value was calculated by comparing the number of S-rand values to *S-obs* delta ELPD for testing a statistically significant pathway enrichment. A higher *S-rand* than *S-obs* indicated a pleiotropic pathway, while a lower S-rand than *S-obs* indicated a causal pathway;

$$P \mathrm{value}=\frac{1}{10^{7}}\sum_{i=1}^{10^{7}} I (|Sum-randi |\geq| Sum-obs|)$$

Since CAUSE works with LD-independent tag SNPs filtered by p-values, we used PLINK to identify SNPs tagged by each tag SNPs (option --show-tags’, *r^2^* > 0.1 in a 250kb window, hg19, 1KG reference panel, CEU sub-population), and these tagged SNPs collectively represent haploblocks. Within each haploblock, each tagged SNP was assigned an LD-weighted ELPD value equal to the tag’s ELPD but shrank by its LD correlation (r^2^) with the tag SNP. Genes intersecting with tagged SNPs inherited the SNP’s LD-weighted ELPD value. This way, neighboring genes to tag SNPs were also considered for pathway analyses. Longer genes incorporate more tagged SNPs, but (*r^2^*) was fall with increasing distances to the tag SNP. Given the hierarchical nature of GO annotations, enriched GO term lists often contain redundant and overlapping terms, making them challenging to summarize and interpret. We adjusted p-values for multiple comparisons by dividing nominal p-value (0.05) to number of clusters using Bonferroni method.

**Supplementary Result 1**

**Cross-sectional analysis of chronic pain and asthma traits**

We found bidirectional associations between chronic pain and asthma. We examined chronic pain, defined as pain persisting ≥ 3 months, through multiple phenotypes assessed across seven distinct body sites, widespread pain, and MCP—a quantitative trait counting the number of body sites with a report of chronic pain. To examine the relationship of increasing pain burden with other variables, we also used an ordinal scale for counts of sites (one, two, three, four, or five-to-seven sites). Asthma cases were stratified by age-of-onset (childhood: <18 years, *N*=12,050; adult: 18-40 years, *N*=11,176 ; and late: >40 years, *N*=15,811; controls *N*=363,681). This yielded nine chronic pain phenotypes (plus five ordinal categories) and three asthma phenotypes.

Female sex increased risk across pain phenotypes for adult and late asthma but decreased childhood asthma risk (*Supplementary Table 1*). Higher age at recruitment increased the risk for late asthma with an opposite trend for childhood and adult asthma (**Supplementary Tables 1, 2**). Sensitivity analyses excluding subjects with chronic obstructive pulmonary disease (COPD) yielded similar chronic pain association estimates with asthma across phenotypes (**Supplementary Tables 3**).

**Supplementary Result 2**

**Genome-wide association scans**

MCP exhibited the highest number of significant loci among pain traits (48 loci: 24 shared with body-site traits and 24 MCP-specific), followed by headache (29 loci), knee, back, neck, and shoulder pain (21–27 loci), hip pain (9 loci), stomach and abdomen pain (3 loci), and facial and widespread pain, each with 1 locus. We considered prioritized genes for each locus based on location and linkage disequilibrium (LD) of the lead SNP. Notably, the single locus for widespread pain (*VPS54*) was shared with MCP and two single loci on chromosomes 4 (*SLC39A8*) and 18 (*DCC*) linked MCP with four and three musculoskeletal pain traits respectively (**Supplementary Fig. 2**). Among asthma traits, childhood asthma revealed the most loci (70 total), sharing seven with adult asthma. Furthermore, five loci were shared across the three asthma traits, comprising 13 genes. All genes displayed immune function including interleukin family genes (*IL1R2, IL1RL1,* and *IL13*). The number of independent significant SNPs decreased with increasing age-of-onset strata for asthma: childhood (586), for adult (78), and late asthma (19); and for pain traits: MCP (139), headache (122), neck/shoulder (50), knee pain (40), back pain (39), hip pain (16), stomach/abdominal (5), facial pain (1), and widespread pain (1).

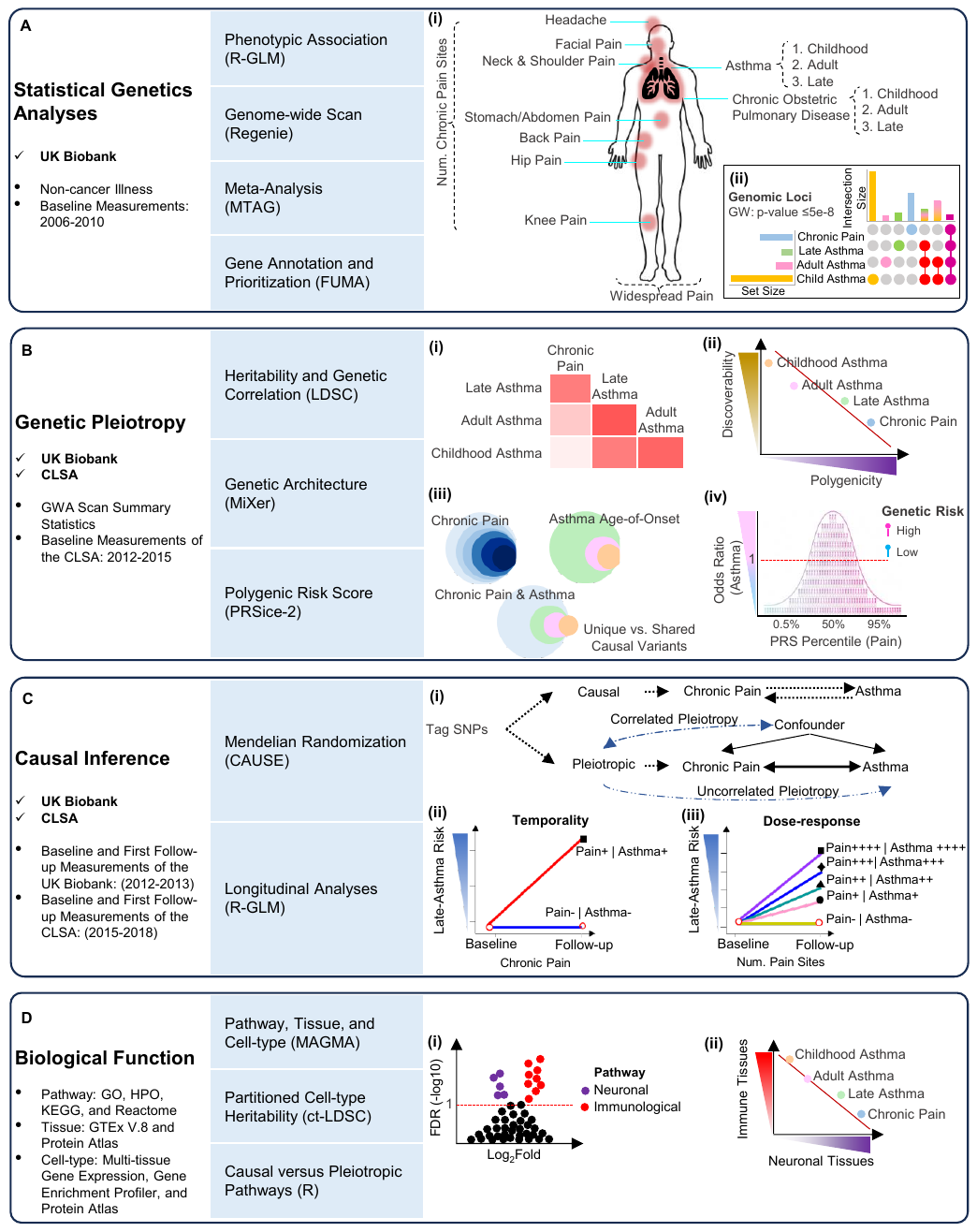
**Supplementary Figure 1.** Overview of the study. **(A)** Data sources used for statistical genetic analyses, including chronic pain traits, asthma and chronic obstetric pulmonary disease (COPD) stratified by age-of-onset. (**i**) Phenotypes were collected at baseline and imaging visits. **(ii)** Genotype and imputed data were used for GWA scans, and significant SNPs from trait-specific GWA scans and meta-analyses were functionally annotated. **(B)** Genetic pleiotropy between chronic pain and asthma was examined using **(i)** genetic correlation, **(ii-iii)** univariate and bivariate MiXeR models, and **(iv)** cross-trait/cross-population polygenic risk score (PRS) analyses. **(C)** The causal modeling framework included **(i)** genome-wide Mendelian Randomization using CAUSE, **(ii)** longitudinal analysis of late asthma incidence at follow-up based on baseline chronic pain status, and **(iii)** a dose-response relationship between the number of chronic pain sites and late asthma risk. Phenotypes were collected at baseline and first follow-up visits **(D)** Biological interpretation involved **(i-ii)** gene-set and tissue-specific enrichment analyses, gene-property and cell-type heritability enrichment.

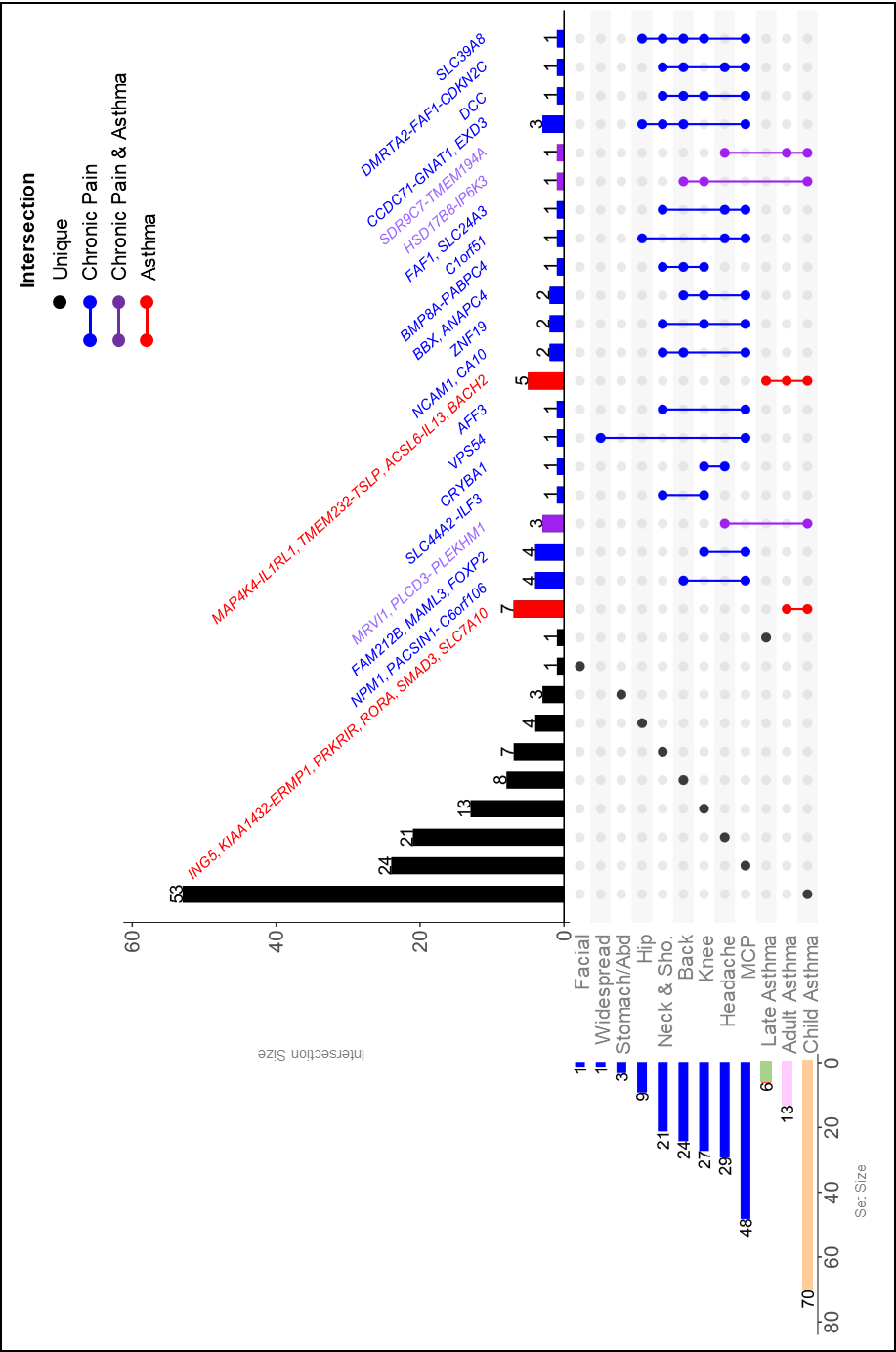

**Supplementary Figure 2.** Shared loci across chronic pain and asthma traits based on trait specific GWA scans. UpSet plot illustrating the number of genomic loci shared across chronic pain traits and asthma age-of-onset strata. The intersection matrix displays unique and overlapping loci and mapped genes, with bars above indicating total loci per combination. The side histogram shows total loci per trait, with colored bars denoting shared loci and black bars indicating trait-specific loci. Vertical lines highlight intersections within chronic pain traits (blue), within asthma strata (red), and between chronic pain and asthma traits (purple).

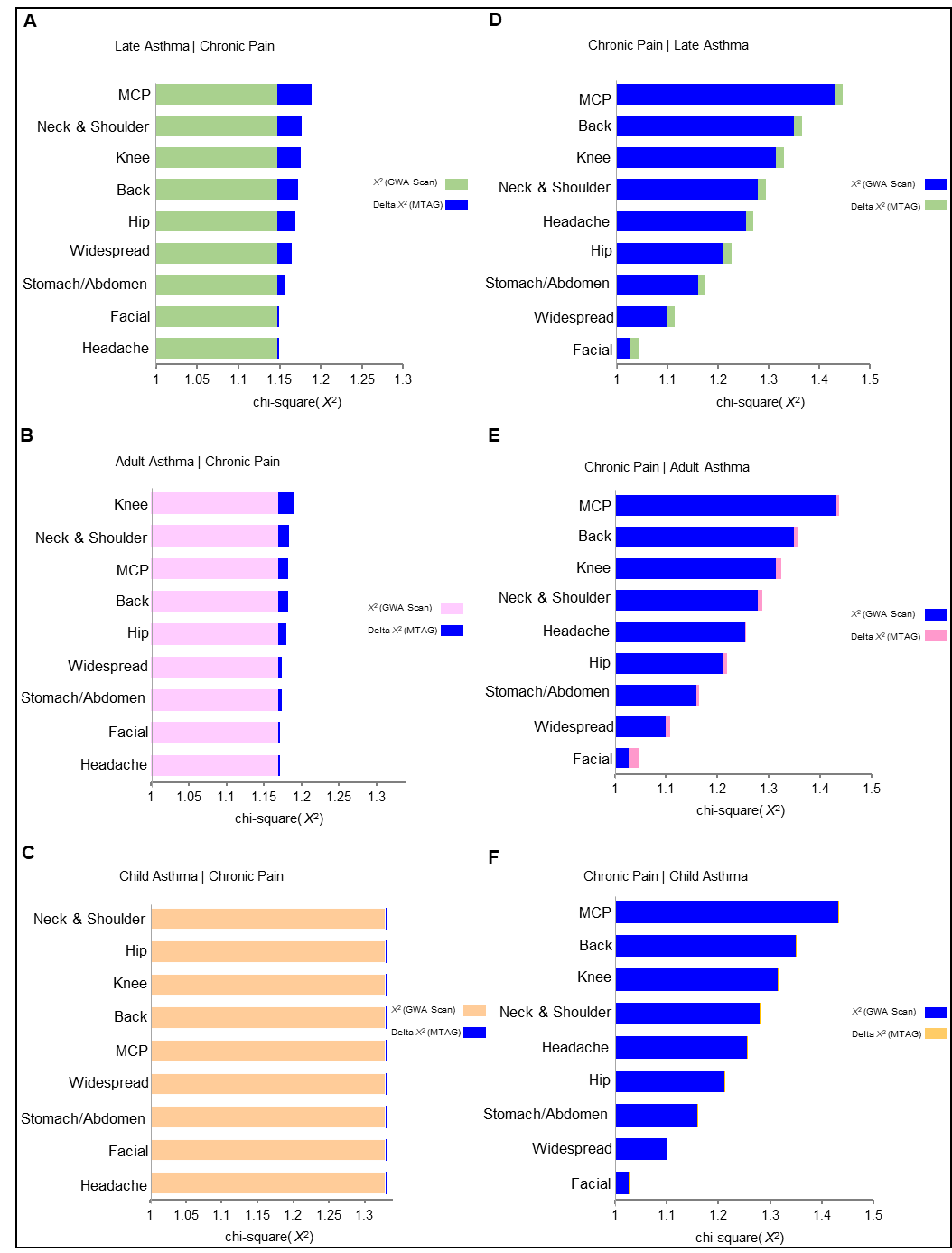

**Supplementary Figure 3.** SNP-level power gain (delta χ²) from cross-trait meta-analysis of asthma and chronic pain traits. Bar plots show the increase in SNP effect size statistics (delta χ²) from meta-analysis relative to trait-specific GWA scans. **(A–C)** Meta-analysis boosting asthma strata with chronic pain traits. **(D–F)** Meta-analysis boosting chronic pain traits with asthma strata. Delta χ² reflects the change in chi-squared values (χ²) for SNPs between the original and boosted analyses.

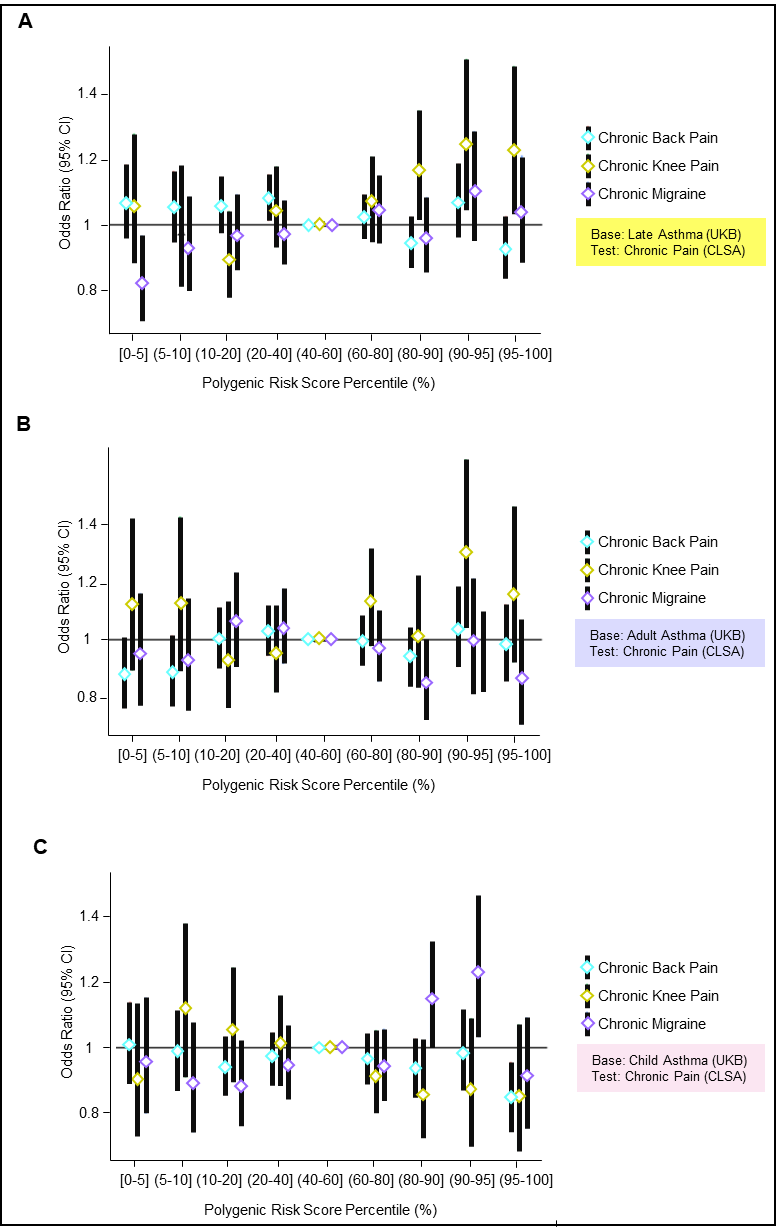

**Supplementary Figure 4.** PRS-based prediction of chronic pain risk in CLSA using asthma age-of-onset traits from the UKB. **(A)** PRS for late asthma. **(B)** PRS for adult asthma. **(C)** PRS for childhood asthma. Odds ratios (ORs) for chronic pain are shown across polygenic risk score (PRS) percentile bins (<5% to >95%), with PRS constructed from asthma strata in the UK Biobank and applied to the Canadian Longitudinal Study on Aging (CLSA). Horizontal black lines indicate no association (OR = 1); vertical black lines represent 95% confidence intervals.

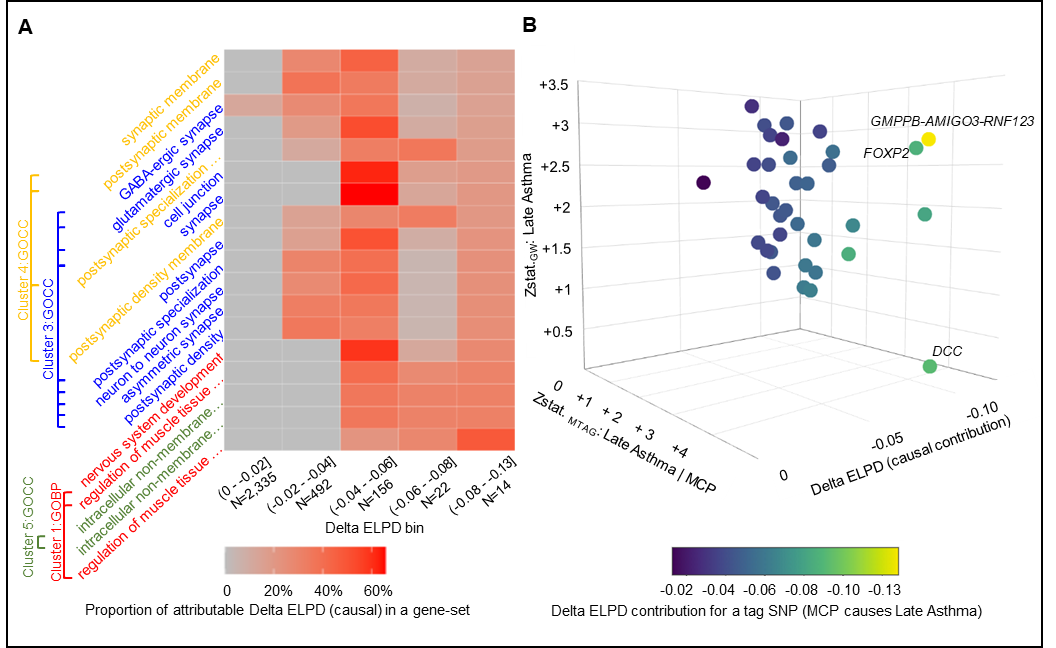

**Supplementary Figure 5. Causal gene function and brain structure associations for MCP and late asthma.** **(A)** Classification of causal gene-set contributions using Mendelian randomization (MR) in the CAUSE framework, grouped into bins by effect size. **(B)** 3D scatter plot showing causal gene signal strength from trait-specific GWA scan Z-statistics, MTAG Z-statistics, and causal contribution (Delta ELPD) from the MR model.
